# Cumulative COVID-19 incidence, mortality, and prognosis in cancer survivors: a population-based study in Reggio Emilia, Northern Italy

**DOI:** 10.1101/2020.11.18.20233833

**Authors:** Lucia Mangone, Francesco Gioia, Pamela Mancuso, Isabella Bisceglia, Marta Ottone, Massimo Vicentini, Carmine Pinto, Paolo Giorgi Rossi

## Abstract

The aim of this population-based study was to evaluate the impact of having had cancer on COVID-19 risk and prognosis during the first wave of the pandemic (27 February – 13 May 2020) in Reggio Emilia Province. Prevalent cancer cases diagnosed between 1996 and December 2019 were linked with the provincial COVID-19 surveillance system. We compared cancer survivors (CS)’ cumulative incidence of being tested, testing positive for SARS-CoV-2, being hospitalized, and dying of COVID-19 with that of the general population; we compared COVID-19 prognosis in CS and in patients without cancer.

15,391 people (1527 CS) underwent RT-PCR for SARS-CoV-2, of whom 4541 (447 CS) tested positive; 541 (113 CS) died of COVID-19. The cumulative incidences of being tested, testing positive, COVID-19 hospitalization, and death were lower in CS: age- and sex-adjusted incidence rate ratios were 1.28 [95%CI = 1.21, 1.35], 1.06 [95%CI = 0.96, 1.18], 1.27 [95%CI = 1.09, 1.48], and 1.39 [95%CI = 1.12, 1.71], respectively. CS had worse prognosis when diagnosed with COVID-19, particularly those below the age of 70 (age- and sex-adjusted odds ratio (OR) of death 5.03; [95%CI = 2.59, 9.75]), while the OR decreased after age 70. The OR of death was higher for patients with a recent diagnosis, i.e. <2 years (OR=2.92; 95%CI = 1.64, 5.21), or metastases (OR=2.09; 95%CI = 0. 88, 4.93).

Cancer patients showed the same probability of being infected, despite a slightly higher probability of being tested, than the general population, nevertheless they were at higher risk of death once infected.

**Novelty and impact:** Cancer survivors during the first wave of the pandemic showed higher COVID-19 cumulative incidence and mortality. When infected, they had worse prognosis, particularly in people younger than age 70 or those with a recent diagnosis.

## Introduction

After China, Italy was one of the first countries that experienced a tragic increase in the incidence of and mortality from COVID-19, with more than 232,000 cases and over 33,000 deaths by the end of May 2020.^1^

The first Chinese study, on a very small group of 18 patients, showed that cancer patients presented a higher risk of SARS-CoV-2 infection and a higher risk of requiring mechanical ventilation or Intensive Care Unit (ICU) admission compared to the general population.^2^

Further studies have found that COVID-19 patients with cancer tend to have much more severe symptoms and a nearly 3-fold higher death rate than COVID-19 patients without cancer.^3^ Especially hematologic, lung, and metastatic cancer patients demonstrated higher rates of severe events compared with patients without cancer.^3^ In addition, patients who underwent cancer surgery and later contracted SARS-CoV-2 showed higher death rates and higher chances of having critical symptoms.^4,5^ Thus, several authors have suggested that immunosuppressed status (whether caused by the disease itself or by treatment) of some cancer patients increases their risk of SARS-CoV-2 infection and of worse outcomes compared with the general population.^6,7^

In Italy, over 377,000 new cases of cancer are estimated to have occurred in 2020, with an estimate of about 3.6 million cancer survivors; this prevalence is estimated to increase dramatically in the next few years.^8^

The aim of this work was to evaluate the impact of having had cancer on COVID-19 risk and prognosis in an Italian province with a high incidence rate of tumors and which also saw a high cumulative incidence of COVID-19 between late February and early May 2020. To achieve this aim, we compared the risk of undergoing a SARS-CoV-2 test, testing positive, being hospitalized, and dying of COVID-19 of people diagnosed with cancer in the previous 25 years with the same risks of the general population. Considering all COVID-19 patients, we also compared cancer survivors’ risk of dying of COVID-19 with that of other COVID-19 patients.

## Methods

### Study design

This was a population-based cohort study using registry data of the Reggio Emilia Province.

### Setting

Reggio Emilia Province, in northern Italy, has a population of about 532,000 inhabitants. The Local Health Authority, the local public entity of the Italian National Health Service, provides hospital, outpatient, primary, and preventive care to the entire population residing in the province.

With the spread of the SARS-CoV-2 virus in northern Italy, the following measures were adopted in Reggio Emilia Province: on February 22, schools were closed, and social restrictions were imposed; on 8 March, mobility and travel restrictions were imposed, and on 11 March, only essential services were permitted to remain open. During this phase, all cases with suspicious symptoms (fever, cough, dyspnea) were tested. The first case of SARS-CoV-2 disease (COVID-19) in the province was diagnosed on February 27, 2020. Up to May 13, the end of the study recruitment period, there were more than 4500 confirmed cases in the province; the epidemic was still spreading, but at a slower rate, and cumulative incidence reached about 9 per 1000.

### Participants

This study included all the residents in Reggio Emilia Province on December 31, 2019. Information on SARS-CoV-2 tests, COVID-19 patients, and prevalent cancer patients was linked. A COVID-19 patient was diagnosed based on real-time reverse transcription polymerase chain reaction (RT-PCR) of nasopharyngeal specimens according to WHO indications; a cancer patient, excluding nonmelanoma skin cancer and brain (non-malignant) cancer, was defined as such if present in the Reggio Emilia Cancer Registry (RE-CR) archive and with a malignant cancer.

### Data sources

The Reggio Emilia Cancer Registry registers all new cancer diagnoses occurring in people residing in the Reggio Emilia Province. The main information sources of the RE-CR are the anatomic pathology reports, the hospital discharge records, and mortality data. The RE-CR, which covers all the resident population in the Reggio Emilia Province, has been active since 2000 and has registered all incident cases from 1996 to 2019, with active follow-up for deaths and residence of all prevalent cases updated to 31 December 2019. It collects information on site, morphology, partial staging (presence of metastases), mode of diagnosis, and survival.^9^

All RT-PCR SARS-CoV-2 tests performed in Italy must be recorded in the national case-based integrated COVID-19 surveillance system.^10^ This surveillance system contains data on all COVID-19 patients, collected by the Public Health Department of the Local Health Authority during epidemiologic investigations through: 1) daily reports from COVID-19 labs for all positive RT-PCR tests; 2) an initial epidemiologic investigation conducted through phone interviews with all cases, followed by daily phone calls to patients cared for in outpatient settings, both conducted by the Public Health Department of each Local Health Authority; 3) daily reports extracted from electronic medical records for hospitalized patients; 4) check of death records to assess mortality, particularly in outpatient settings.

The whole list of cases tested for SARS-CoV-2 (positive and negative) from late February up to 13 May 2020 was linked with the entire archive of prevalent cases of malignant cancer on 31 December 2019 present in the RE-CR database.

### Outcome measures

From the COVID-19 surveillance system data, we report the cumulative incidence up to 13 May 2020 of: a) being tested for SARS-CoV-2; b) testing positive for SARS-CoV-2 (according to the COVID-19 definition adopted by the Italian Ministry of Health, these are all COVID-19 cases); c) hospitalizations of COVID-19 patients; d) deaths among COVID-19 patients. All COVID-19 cases were followed up for 45 days from the first positive test for death or hospitalization. Furthermore, hospital admissions occurring in the 8 days before SARS-CoV-2 positivity were considered as COVID-19 hospitalizations. Follow-up was closed on 28 June 2020.

Based on the data collected by the Reggio Emilia Cancer Registry, all the tested subjects were grouped according to their oncological history: patients who had never been diagnosed with a malignancy *versus* patients with a clinical history of malignancy at any time since 1996. Cancer patients were then categorized according to the time since cancer diagnosis (<2 years, 2-5 years, and >5 years), cancer site, and presence of metastases at diagnosis. Age was calculated on 31 December 2019.

### Statistical analysis

Pearson’s chi-square test was used to examine differences in the proportions of subjects with and without cancer and tested/not tested for SARS-CoV-2. We report age- and sex-adjusted incidence rate ratios, with relative 95% confidence intervals (95% CI) using Poisson regression, for cumulative incidences. The outcomes of interest for this analysis were: cancer survivors’ being tested for SARS-CoV-2, having a positive test, being hospitalized, and dying of COVID-19, compared with the same outcomes of those who had never had a cancer diagnosis. Multivariable analysis was performed using a logistic regression model to measure the odd ratios, with relative 95% CI, of hospitalization and death for COVID-19 patients with cancer, adjusting for age and sex. STATA v.14.1 (StataCorp LP 4905 Lakeway Drive, Texas 77845 USA) was used for all analyses.

## Results

On 31 December 2019, there were 27,386 residents of the Reggio Emilia Province still alive after a cancer diagnosis, for a prevalence of 5.1%.

Between 27 February and 13 May 2020, 15,391 residents in the Reggio Emilia Province underwent molecular testing for SARS-CoV-2 (2.9% of the resident population); testing was more frequent among the elderly and among females; 1527 cancer survivors underwent testing (5.6% of total cancer survivors) (Table 1). The age- and sex-adjusted cumulative incidence ratio of being tested was 1.28 (95% CI = 1.21, 1.35) (Table 2).

**Table 1.**
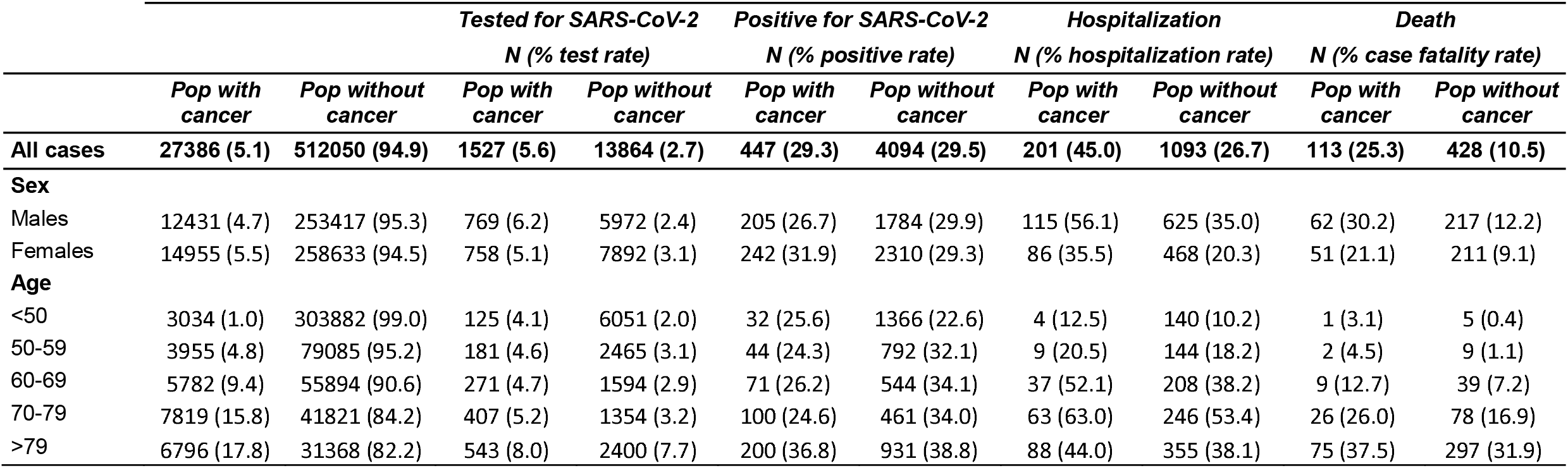
Distribution of 15,391 patients who tested for SARS-CoV-2, were positive for SARS-CoV-2, COVID-19 hospitalization, and death, by sex, age, and cancer history.

**Table 2.**
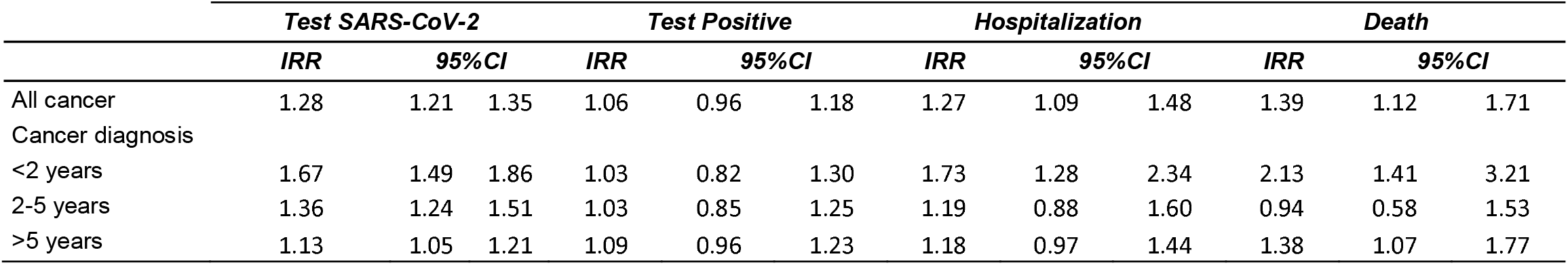
Incidence rate ratio (IRR) of being tested, having a positive test, being hospitalized, and dying within 45 days from COVID-19 diagnosis for people with previous diagnosis of cancer compared with the general population without cancer.

|  | *Test SARS-CoV-2* |  |  | *Test Positive* |  |  | *Hospitalization* |  |  | *Death* |  |  |
| --- | --- | --- | --- | --- | --- | --- | --- | --- | --- | --- | --- | --- |
|  | *IRR* | *95%CI* |  | *IRR* | *95%CI* |  | *IRR* | *95%CI* |  | *IRR* | *95%CI* |  |
| All cancer | 1.28 | 1.21 | 1.35 | 1.06 | 0.96 | 1.18 | 1.27 | 1.09 | 1.48 | 1.39 | 1.12 | 1.71 |
| Cancer diagnosis |  |  |  |  |  |  |  |  |  |  |  |  |
| <2 years | 1.67 | 1.49 | 1.86 | 1.03 | 0.82 | 1.30 | 1.73 | 1.28 | 2.34 | 2.13 | 1.41 | 3.21 |
| 2-5 years | 1.36 | 1.24 | 1.51 | 1.03 | 0.85 | 1.25 | 1.19 | 0.88 | 1.60 | 0.94 | 0.58 | 1.53 |
| >5 years | 1.13 | 1.05 | 1.21 | 1.09 | 0.96 | 1.23 | 1.18 | 0.97 | 1.44 | 1.38 | 1.07 | 1.77 |

Of the 15,391 tests performed, 4541 (29.5%) were positive for SARS-CoV-2, i.e. were COVID-19 cases, with no differences between those with a cancer diagnosis (29.3%) and those without (29.5%) (Table 1). The cumulative incidence of SARS-CoV-2-positive tests was 0.8% in the general population and 1.6% in people with cancer (Table 1). The age- and sex-adjusted COVID-19 incidence ratio was 1.06 (95% CI = 0.96, 1.18) (Table 2).

The overall cumulative incidence of hospitalizations was 0.24%, higher in males and in older people; in cancer patients it was 0.7% (201 hospitalized cases) (Table 1), corresponding to an age- and sex-adjusted hospitalization incidence rate ratio of 1.27 (95% CI = 1.09, 1.48) (Table 2). The overall cumulative mortality was 0.1% in the general population, reaching 1.0% in people over age 80. Crude cumulative mortality was higher in people with cancer (113 deaths, 0.41%), while the age- and sex-standardized mortality incidence rate ratio was 1.39 (95% CI = 1.12, 1.71) (Table 2).

The multivariable logistic analysis (Table 3) among COVID-19-positive patients showed that females had a lower risk of being hospitalized and of dying (OR 0.43; 95% CI = 0.37, 0.49 and OR 0.48; 95% CI = 0.39-0.59, respectively). People ages 70-80 had the highest probability of being hospitalized, while those age <70 had a lower probability than people age >80. The probability of death increased with age, with ORs for those below age 70 compared to those over 80 of 0.04 (95% CI = 0.03, 0.05) and for age 70-80 *vs* over 80 of 0.37 (95% CI = 0.29, 0.48). We observed a strong interaction between age and cancer in the effect on fatality rate and, to a lesser degree, also on hospitalization. The OR of being hospitalized for COVID-19 among those below age 70 with a previous diagnosis of cancer was 2.6 (95% CI = 1.8, 3.7), which decreased with age (among those over age 80, OR 1.2 (95% CI = 0.8, 1.6). The OR of dying was 5.0 (95% CI = 2.6, 9.8) for patients below age 70, which decreased with age (OR 1.1; 95% CI = 0.8, 1.6 for patients over age 80).

**Table 3.** Multivariable logistic regression of positive cases by age and sex, and age-stratified multivariable logistic regression for cancer history.

|  | Hospitalization |  |  | Death |  |  |
| --- | --- | --- | --- | --- | --- | --- |
|  | OR* | 95%CI |  | OR* | 95%CI |  |
| Sex |  |  |  |  |  |  |
| Male | 1 |  |  | 1 |  |  |
| Female | 0.43 | 0.37 | 0.49 | 0.48 | 0.39 | 0.59 |
| Age |  |  |  |  |  |  |
| < 70 | 0.33 | 0.28 | 0.38 | 0.04 | 0.03 | 0.05 |
| 70 - 80 | 1.62 | 1.32 | 1.98 | 0.37 | 0.29 | 0.48 |
| > 80 | 1 |  |  | 1 |  |  |
| Cancer and age |  |  |  |  |  |  |
| Age<70 |  |  |  |  |  |  |
| Without cancer | 1 |  |  | 1 |  |  |
| With cancer | 2.58 | 1.79 | 3.72 | 5.03 | 2.59 | 9.75 |
| Age 70-80 |  |  |  |  |  |  |
| Without cancer | 1 |  |  | 1 |  |  |
| With cancer | 1.37 | 0.92 | 2.06 | 1.90 | 1.20 | 3.01 |
| Age>80 |  |  |  |  |  |  |
| Without cancer | 1 |  |  | 1 |  |  |
| With cancer | 1.15 | 0.82 | 1.61 | 1.13 | 0.80 | 1.58 |

Excess mortality in cancer patients was higher in the presence of metastases, but differences could be due to chance (OR 2.1; 95% CI = 0.9, 4.9) (Table 4). Concerning timing, the strongest mortality excess was for cancers diagnosed less than two years before the onset of COVID-19 symptoms (OR 2.9; 95% CI = 1.6, 5.2). The patterns were similar for hospitalization, except for 2-5 years from cancer diagnosis. We also analyzed the distribution by cancer site: among the 447 SARS-CoV-2-positive patients with cancer, the sites most frequently involved were breast (101 cases), digestive organs (87 cases, of which 46 colon, 22 rectum, 13 stomach, 3 pancreas, 2 liver, 1 biliary tract), urinary tract (54 cases, of which 38 bladder, 14 kidney, 2 upper urinary tract), and male genital organs (58 cases, of which 53 prostate, 3 testis, 1 penis, and 1 other and undefined male genital organs). COVID-19 patients with cancer of the gastrointestinal tract, lymphoma, or other hematological neoplasms showed the greatest excess of hospitalizations; cancer of the urinary tract, other hematological neoplasms, melanoma, and female genital organs showed the strongest excess mortality.

**Table 4.**
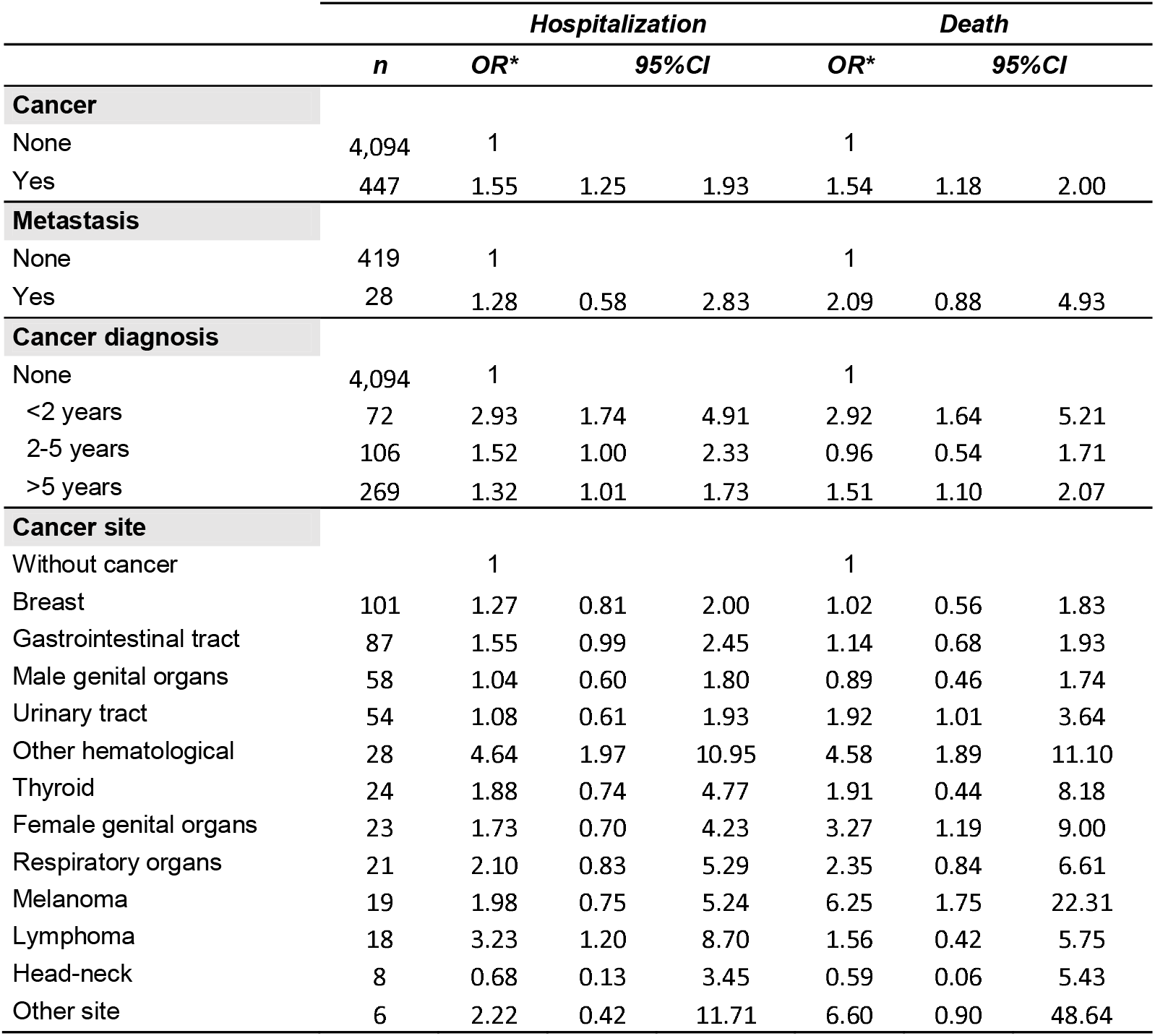
Multivariable logistic regression of positive cases related to clinical outcomes, adjusted for age and sex.

## Discussion

Although people with a previous diagnosis of cancer had a slightly higher probability of being tested for SARS-CoV-2 than the general population, the risk of infection was comparable with that of the general population without cancer. On the other hand, the risk of COVID-19-related hospitalization and death were higher among cancer patients than in the general population without cancer. The higher probability of being tested, but not of infection, was also observed in patients with a recent infection. In this group the excess risk of being tested was much higher than in patients with older diagnoses, probably because these patients had more frequent access to health services. This suggests that the control measures adopted in oncology and diagnostic follow-up clinics have been effective in not increasing the risk of infections in these patients, but also that cancer survivors are not more susceptible to infections, or that any increased susceptibility has been counterbalanced by protective measures. In fact, it must be taken into consideration that the Italian Government implemented special protection measures for people with chronic diseases, including cancer, such as exemption from any work not allowing social distancing.

Once having COVID-19, the probabilities of being hospitalized and of dying were much higher for people with a previous diagnosis of cancer that were below the age of 70; excess mortality was appreciable only up to the age of 80, while after 80 the excess was compatible with random fluctuation. For hospitalization, the excess decreased with age.

Higher fatality and hospitalization rates for COVID-19 patients with a previous diagnosis of cancer have been observed in several studies,^11-17^ including a recent large registry-based Italian study.^11^Instead, we did not find any studies comparing the risk of SARS-CoV-2 infection and COVID-19 death in cancer survivors with that in the general population. Only a few case series have tried to infer the risk of infection, comparing the prevalence of SARS-CoV-2 infection in cancer patients with that in health care providers^12^ or with that in the general population,^13-15^ or comparing the prevalence of cancer among COVID-19 patients with that in the general population.^16^ These studies have provided inconsistent results.^12,17^

Not surprisingly, the proportion of people with a previous cancer diagnosis among COVID-19 patients (about 9.8% in our province) was much higher than that reported in other studies from China, ranging between 2.7%^18^ and 1%.^2^

In our cohort of COVID-19 patients, a previous diagnosis of cancer increased the risk of death up to the age of 80, after which the impact of having had cancer was modest or null; the decreasing impact on prognosis could be partially due to a higher proportion of cured cancer survivors, for whom we would expect a small or null impact in the age group >80. In fact, the proportion of patients who had a cancer diagnosis more than 5 years earlier was 70.4% and 51.7% in patients over 80 and below 70, respectively (Supplementary Table). A systematic review including about 50,000 COVID-19 patients observed the same phenomenon: the impact of previous cancer on survival was important before the age of 65 but was almost null over that age.^19^ This has also been observed for the impact of all comorbidities on COVID-19 survival, which decreases as age increases.^10^

It is worth noting that the excess risks of death and hospitalization were stronger in patients with a recent diagnosis of cancer and in patients with metastases at diagnosis. This is consistent with the findings of Liang and colleagues,^2^ who found an excess only in patients who had recently received treatment, and with the results of Dai and colleagues, who reported an excess for patients who had had a diagnosis of stage IV cancer.^3^ Data from a large cohort of patients with active cancer and COVID-19 in the UK did not confirm an excess risk due to recently undergoing chemotherapy or radiotherapy compared to the patients that did not. The excess risk of those with metastases, however, was confirmed.^20^

Thus, it remains unclear whether the increased risk of death was due to the direct effects of cancer or to treatment (surgery and chemotherapy). However, our data, plus insights from other studies, suggest that the magnitude of the effect on COVID-19 prognosis is greater during the active phase of cancer, after which the effect decreases.

The strongest excess mortality was observed in patients with cancer of the female genital organs, urinary tract, other hematologic neoplasms, and melanoma, while for lymphomas and gastrointestinal tract (borderline), we observed a considerable excess of hospitalizations but just a small excess of deaths. It is not easy to compare our results with those of other studies because most included only hospitalized patients; if the risk of hospitalization does not reflect that of death, relative risk among hospitalized patients is affected by a collider bias. Nevertheless, our data are not consistent with those of the large UK cohort, where small excess risk was observed for lymphoma and respiratory cancers.^19^

These results make it difficult to put together a profile of patients at higher risk of dying of COVID-19 based on cancer site and phase of care, for whom particular measures, including delaying treatments, could be taken to reduce the risk of infection, as suggested by some authors.^21,22^ The only feature that clearly emerged from our findings was recent diagnosis and the presence of metastases.

Our data suggest that public awareness and measures allowing cancer survivors to control their risk of infection resulted in their risk being comparable to that of the general population even when they were in an active phase of care or follow-up. This is an important message for the policy makers, physicians, and patients that are trying to better manage cancer during this public health emergency.^23^

The main strength of this study is its population-based design, which eliminates any selection bias occurring in case series. Furthermore, the assessment of exposure, i.e. previous diagnosis of cancer, was conducted through the linkage with a cancer registry with 25 years of prevalence data and timely registration of incident cases (to 31 December 2019). It is worth noting that this information was acquired before the onset of the pandemic and is thus completely independent of outcome occurrence. The main limitation of our study is that we do not have any information on treatment or on comorbidities, which could have influenced outcomes. Furthermore, because we could not include cancer patients with a diagnosis occurring in 2020, we could not observe the phase of diagnosis and disease assessment, which for many cancer sites is very intensive in terms of access to healthcare facilities.

## Conclusion

Our population-based study showed that during the peak of the COVID-19 epidemic in northern Italy, cancer survivors had similar cumulative incidence of COVID-19, despite a slightly higher probability of being tested. On the other hand, they had a greater risk related to hospitalizations and death once infected, especially in the age group <70 years or those with a recent diagnosis.

## Data Availability

Researchers who would like to access individual data should present their request, together with a study protocol, to the Area Vasta Emilia Nord Ethics Committee for approval.

## Ethics statement

The study was approved by the Area Vasta Emilia Nord Ethics Committee (no. 2020/0045199). The Ethics Committee authorized the use of patient data, even in the absence of consent, if all reasonable efforts had been made to contact that patient.

## Conflicts of interest

The authors have no conflicts of interest to disclose.

## List of abbreviations

CI: Confidence Interval
COVID-19: Coronavirus Disease 19
CS: Cancer Survivors
ICU: Intensive Care Unit
IRR: Incidence Rate Ratios
OR: Odds Ratio
RE-CR: Reggio Emilia Cancer Registry
RT-PCR: Real-time Polymerase Chain Reaction
SARS-CoV-2: Severe Acute Respiratory Syndrome Coronavirus 2
UK: United Kingdom
WHO: World Health Organization

## Legend of tables

Supplementary Table. Distribution of outcomes by age and cancer diagnosis in cancer population positive for SARS-Cov-2.

## Notes

### Competing Interest Statement

The authors have declared no competing interest.

### Funding Statement

No external funding were received.

## References

1. Dong E, Du H, Gardner L. An interactive web-based dashboard to track COVID-19 in real time. Lancet Infect Dis. 2020; 20: 533–534.

2. Liang W, Guan W, Chen R, Wang W, Li J, Xu K, Li C, Ai O, Lu W, Liang H, Li S, He J. Cancer patients in SARS-CoV-2 infection: a nationwide analysis in China. Lancet Oncol. 2020; 21: 335–337.

3. Dai M, Liu D, Liu M, Zhou F, Li G, Chen Z, Zhang Z, You H, Wu M, Zheng Q, Xiong Y, Xiong H, Wang C, Chen C, Xiong F, Zhang Y, Peng Y, Ge S, Zhen B, Yu T, Wang L, Wang H, Liu Y, Chen Y, Mei J, Gao X, Li Z, Gan L, He C, Li Z, Shi Y, Qi Y, Yang J, Tenen D, Chai L, Mucci LA, Santillana MH. Patients with Cancer Appear More Vulnerable to SARS-COV-2: A Multicenter Study during the COVID-19 Outbreak. Cancer Discov. 2020; 10:783–791.

4. Wu Z, McGoogan JM. Characteristics of and important lessons from the coronavirus disease 2019 (COVID-19) outbreak in China: summary of a report of 72314 cases from the Chinese Center for Disease Control and Prevention. JAMA 2020; 323:1239–42.

5. Gosain R, Abdou Y, Singh A, Rana N, Puzanov I, Ernstoff MS. COVID-19 and Cancer: a Comprehensive Review. Curr Oncol Rep 2020; 22:53.

6. International Pharmaceutical Federation. Coronavirus SARS-CoV-2 outbreak: information and guidelines for pharmacists and the pharmacy workforce. Febuary 12, 2020.

7. Zhang L, Zhu F, Xie L, Wang C, Wang J, Chen R, Jia P, Guan HQ, Peng L, Chen Y, Peng P, Zhang P, Chu Q, Shen Q, Wang Y, Xu SY, Zhao JP, Zhou M. Clinical characteristics of COVID-19-infected cancer patients: a retrospective case study in three hospitals within Wuhan, China. Ann Oncol. 2020; 31:894–901.

8. I numeri del cancro in Italia 2020. AIOM-AIRTUM-SIAPEC-IAP. Intermedia Editore, October 2020.

9. Cancer Incidence in Five Continents, Vol. XI (electronic version). Lyon: International Agency for Research on Cancer. Available from: https://ci5.iarc.fr, xaccessed [23th October 2020].

10. Ferroni E, Giorgi Rossi P, Spila Alegiani S, Trifirò G, Pitter G, Leoni O, Cereda D, Marino M, Pellizzari M, Fabiani M, Riccardo F, Sultana J, Massari M; ITA-COVID Working Group. Survival of Hospitalized COVID-19 Patients in Northern Italy: A Population-Based Cohort Study by the ITA-COVID-19 Network. Clin Epidemiol 2020; 121337–1346.

11. Rugge, M., Zorzi, M. & Guzzinati, S. SARS-CoV-2 infection in the Italian Veneto region: adverse outcomes in patients with cancer. Nat Cancer 2020; 1:784–788.

12. He W, Chen L, Chen L, Yuan G, Fang Y, Chen W, Wu D, Liang B, Lu X, Ma Y, Li L, Wang H, Chen Z, Li Q, Gale RP. COVID-19 in persons with haematological cancers. Leukemia 2020; 34:1637–45.

13. van Dam PA, Huizing M, Mestach G, Dierckxsens S, Tjalma W, Trinh XB, Papadimitriou K, Altintas S, Vermorken J, Vulsteke C, Janssens A, Berneman Z, Prenen H, Meuris L, Vanden Berghe W, Smits E, Peeters M. SARS-CoV-2 and cancer: Are they really partners in crime? Cancer Treat Rev. 2020; 89:102068.

14. Yu J, Ouyang W, Chua MLK, Xie C. SARS-CoV-2 Transmission in Patients With Cancer at a Tertiary Care Hospital in Wuhan, China. JAMA Oncol 2020; 25:e200980.

15. Barlesi F, Foulon S, Bayle A, Gachot B, Pommeret F, Willekens C, Stoclin A, Merad M, GriscelliI F, Micol JB, Sun R, Nihouarn T, Balleygier C, André F, Scotte F, Besse B, Soria JC, Albiges L, Roussy G. Outcome of cancer patients infected with COVID-19, including toxicity of cancer research. Presented at: 2020 virtual annual meeting of the American Association for Cancer Research; April 27–28; 2020.

16. El Gohary GM, Hashmi S, Styczynski S, Kharfan-Dabaja MA, Alblooshi RM, de la Cámara R, Mohmed S, Alshaibani A, Cesaro S, Abd El-Aziz N, Almaghrabi R, Gergis U, Yasser M. The risk and prognosis of COVID-19 infection in cancer patients: A systematic review and meta-analysis. Hematol Oncol Stem Cell Ther. 2020; S1658-3876(20)30122-9.

17. Robinson AG, Gyawali B, Evans G. COVID-19 and cancer: do we really know what we think we know? Nat Rev Clin Oncol 2020; 17:386–8.

18. Ma J, Yin J, Qian Y, Wu Y. Clinical characteristics and prognosis in cancer patients with COVID-19: A single center’s retrospective study. J Infect2020; 81:318–356.

19. Giannakoulis VG, Papoutsi E, Siempos II. Effect of Cancer on Clinical Outcomes of Patients With COVID-19: A Meta-Analysis of Patient Data. JCO Glob Oncol. 2020; 6:799–808.

20. Lee L, Cazier JB, Starkey T, Turnbull CD; UK Coronavirus Cancer Monitoring Project Team, Kerr R, Middleton G. COVID-19 mortality in patients with cancer on chemotherapy or other anticancer treatments: a prospective cohort study. Lancet. 2020; 395:1919–1926.

21. Al-Quteimat OM, Amer AM. The Impact of the COVID-19 Pandemic on Cancer Patients. Am J Clin Oncol. 2020; 43:452–455.

22. Finley C, Prashad A, Camuso N, Daly C, Aprikian A, Ball CG, Bentley J, Charest D, Fata P, Helyer L, O’Connell D, Moloo H, Seely A, Werier J, Zhong T, Earle CC. Guidance for management of cancer surgery during the COVID-19 pandemic. Can J Surg. 2020; 63:S2–S4.

23. Al-Shamsi HO, Alhazzani W, Alhuraiji A, Coomes EA, Chemaly RF, Almuhanna M, Wolff RA, Ibrahim NK, Chua MLK, Hotte SJ, Meyers BM, Elfiki T, Curigliano G, Eng C, Grothey A, Xie C. A Practical Approach to the Management of Cancer Patients During the Novel Coronavirus Disease 2019 (COVID-19) Pandemic: An International Collaborative Group. Oncologist. 2020; 25:e936–e945.

