## Supplemental Table 1 for "Cumulative COVID-19 incidence, mortality, and prognosis in cancer survivors: a population-based study in Reggio Emilia, Northern Italy"

Supplementary Table. Distribution of outcomes by age and cancer diagnosis in cancer population positive for SARS-Cov-2.

|  | **Positive for** | | **Hospitalization** | | **Death** | |
| --- | --- | --- | --- | --- | --- | --- |
|  | **SARS-CoV-2** | |  |  |  |  |
|  | **n** | **%col** | **n** | **%row** | **n** | **%row** |
| **Cancer diagnosis and age** |  |  |  |  |  |  |
| **Age<70** |  |  |  |  |  |  |
| <2 years | 26 | 17.7 | 15 | 57.7 | 5 | 19.2 |
| 2-5 years | 45 | 30.6 | 13 | 28.9 | 2 | 4.4 |
| >5 years | 76 | 51.7 | 22 | 28.9 | 5 | 6.6 |
| **Age 70-80** |  |  |  |  |  |  |
| <2 years | 27 | 22.3 | 19 | 70.4 | 9 | 33.3 |
| 2-5 years | 27 | 22.3 | 21 | 77.8 | 9 | 33.3 |
| >5 years | 67 | 55.4 | 34 | 50.7 | 16 | 23.9 |
| **Age>80** |  |  |  |  |  |  |
| <2 years | 19 | 10.6 | 10 | 52.6 | 10 | 52.6 |
| 2-5 years | 34 | 19.0 | 11 | 32.4 | 6 | 17.6 |
| >5 years | 126 | 70.4 | 56 | 44.4 | 51 | 40.5 |
